# Differential impact of Covid-19 on incidence of diabetes mellitus and cardiovascular diseases in acute, post-acute and long Covid-19: population-based cohort study in the United Kingdom

**DOI:** 10.1101/2021.12.13.21267723

**Authors:** Emma Rezel-Potts, Abdel Douiri, Xiaohui Sun, Phillip J Chowienczyk, Ajay M Shah, Martin C Gulliford

## Abstract

**Objective:** This study aimed to estimate the incidence of new diabetes mellitus (DM) and cardiovascular diseases (CVD) up to one year after Covid-19 compared with matched controls.

**Methods:** A cohort study was conducted using electronic records for 1,473 family practices with a population of 14.9 million. Covid-19 patients without DM or CVD were individually matched with controls and followed up to October 2021. A difference-in-difference analysis estimated the net effect of Covid-19 allowing for baseline differences and covariates.

**Results:** There were 372,816 Covid-19 patients, with 2,935 CVD and 3,139 DM events, and 372,816 matched controls with 1,193 CVD and 1,861 DM events following the index date. Net incidence of DM increased in acute Covid-19 up to four weeks from index date (adjusted rate ratio, RR 1.71, 1.40 to 2.10) and remained elevated in post-acute (five to 12 weeks from index date; RR 1.17, 1.01 to 1.36) and long-Covid-19 (13 to 52 weeks, 1.20, 1.09 to 1.31). Acute Covid-19 was associated with net increased CVD incidence (RR 6.02, 95% confidence interval 4.84 to 7.47) including pulmonary embolism (RR 14.5, 7.72 to 27.4), atrial arrythmias (6.58, 3.78 to 11.4) and venous thromboses (5.44, 3.22 to 9.17). CVD incidence declined in post-acute Covid-19 (1.68, 1.41 to 2.01) and showed no net increase in long Covid-19 (0.95, 0.85 to 1.06).

**Conclusions:** DM incidence remains elevated up to one year following Covid-19. CVD is increased early after Covid-19 mainly from pulmonary embolism, atrial arrhythmias and venous thromboses.

## INTRODUCTION

Covid-19 is increasingly recognised as a multi-system condition.^1^ Infection of the respiratory tract by the SARS-CoV-2 virus triggers host immune responses that may have systemic effects through activation of inflammatory pathways.^2,3^ Covid-19 may trigger a proinflammatory ‘cytokine storm’ with dysregulated immune response, platelet activation, hypercoagulability, endothelial cell dysfunction and thromboembolism affecting diverse systems with potential for end-organ damage.^4^ Acute Covid-19 infection has been associated with new-onset cardiovascular disease (CVD) and diabetes mellitus (DM.) Cardiac manifestations of Covid-19^5,6^ include cardiac injury with elevated troponin levels, heart failure and increased risk of mortality among patients hospitalised with Covid-19.^7,8^ Covid-19 may also be associated with acute myocardial infarction and ischaemic stroke in the first 4 weeks.^9^ New onset hyperglycaemia has been reported in Covid-19 patients and is associated with worse prognosis. Complications of both pre-existing and new onset DM have been observed, including diabetic ketoacidosis and hyperosmolarity.^10-12^ Possible mechanisms appear to be direct pancreatic damage from SARS-CoV-2 and/or the associated systemic inflammatory syndrome with severe Covid-19, as reflected in high levels of interleukin-6 (IL-6) and tumour necrosis factor alpha [TNFα], causing impaired pancreatic insulin secretion^13^ and insulin resistance.^14^

Since the first evidence of Covid-19 emerged in the beginning of 2020, there have been multiple waves of infection. There were in excess of 1,000 deaths per day in the UK during April 2020 and January 2021 with many more suffering illness of varying severity.^15^ The longer-term outcomes of Covid-19 are now receiving increasing attention. A high proportion of patients report experiencing symptoms continuing for more than 4 weeks after initial presentation with Covid-19.^16^ The distinction has been made between ‘acute Covid-19’ in the first four weeks after infection; ‘post-acute’ Covid-19 (or ‘ongoing symptomatic covid-19’), from 5 to 12 weeks after the first infection; and ‘long’ Covid-19 (or ‘post-covid-19 syndrome’) with symptoms persisting more than 12 weeks after infection.^17^ There is concern that cardiovascular and metabolic outcomes may be compromised in the aftermath of Covid-19 infection. However, susceptibility to and severity of Covid-19 infection is known to be associated with cardiometabolic risk. Furthermore, public health restrictions during the height of the pandemic, including ‘lockdowns’ or ‘stay at home’ orders, were associated with profound changes in diet, exercise habits and other health-related behaviours that might have impacted on cardiovascular disease and diabetes in the general population even in the absence of Covid-19 infection. Controlled studies are therefore needed to evaluate the net long-term impacts of Covid-19 infection on cardiovascular and diabetes outcomes after allowing for pre-morbid differences between cases and controls and changes over time in control participants. However, few studies have reported on long-term follow-up for large population-based samples.

Longitudinal datasets from electronic health records offer opportunities to analyse longer-term Covid-19 outcomes. We employed the Clinical Practice Research Datalink (CPRD), a national database of anonymised primary care electronic health records, to identify a cohort of Covid-19 patients, comparing new CVD and DM diagnoses to a matched cohort with no Covid-19 diagnosis. We aimed to estimate the net effect of Covid-19 on cardiometabolic outcomes over periods of four weeks, three months and 12 months, in order to inform research priorities, clinical services and public health interventions that may be required following acute Covid-19 infection.

## METHODS

### Data source and participant selection

The CPRD Aurum is a large database which includes comprehensive medical record data for a total of 1,473 family practices in England with approximately 14.9 million currently registered patients.^18^ The data, which cover approximately 13% of the population of England, are representative of the general population with regards to geographical distribution, deprivation, age and gender.

Data for all patients with a diagnosis of Covid-19 were extracted from the February 2021 release of CPRD Aurum. The date of the first code for Covid-19 was the index date. Since confirmatory testing was not widely available during the initial phase of the pandemic, we included patients with clinical diagnoses of ‘confirmed’ or ‘suspected’ Covid-19. The Covid-19 cohort was compared with a sample of matched patients who were not recorded with a Covid-19 diagnosis up to the case index date. Patients were randomly sampled from the entire registered population of CPRD Aurum March 2021 release, individually matching on age, gender and family practice. Data were updated to the December 2021 release of CPRD Aurum for final analyses, providing follow-up to mid-October 2021. The records of any control patients diagnosed with Covid-19 after the matched case index date were censored seven days before the control Covid-19 diagnosis date. Matched sets that included case or control patients with prevalent CVD or DM, diagnosed within 12 months of the start of record or more than 12 months before the index date, were excluded.

### Outcome Measures

The study outcomes were first CVD and DM diagnoses. CVD diagnoses were grouped into the categories: myocardial infarction and ischaemic heart disease; atrial arrhythmias, including atrial fibrillation and supraventricular tachycardia; heart failure; cardiomyopathy and myocarditis; pulmonary embolism; venous thrombosis; and stroke. DM diagnoses included diagnoses for type 1 and type 2 diabetes mellitus and initiation of oral hypoglycaemic drugs and insulin. Records of HbA1c were evaluated and a second record of HbA1c ≥48mmol/mol was considered diagnostic of diabetes. Patients were considered to have a ‘type 1 DM’ phenotype if they were aged 35 years or less at diagnosis and were prescribed insulin within 91 days of diagnosis.^19^ Mortality was evaluated from the CPRD date of death.

### Covariates

Covariates were defined using data recorded in the study period before the index date. These included smoking status (non-smoker, current smoker, ex-smoker), body mass index (BMI, underweight <18.5 Kg/m^2^, normal weight 18.5-24.9 18.5 Kg/m^2^, overweight 25.0-29.9 18.5 Kg/m^2^, obese ≥30 18.5 Kg/m^2^, and not recorded), systolic blood pressure (SBP) and diastolic blood pressure (DBP) in categories of 10 mm Hg. Ethnicity was classified as ‘white’, ‘black’, ‘Asian’, ‘mixed’, ‘other’ and ‘not known’. We evaluated the comorbidities of the Charlson comorbidity index as present or absent before the index date and calculated the Charlson score, with a maximum of four or more. ^20 21^ Case and control patients were matched on gender, year of birth and family practice, with the latter offering control for area measures including level of deprivation. Prescriptions for glucocorticoids were evaluated in patients diagnosed with DM.

### Analysis

Follow-up time was divided into periods of 28 days, from one year before the index date to one year after the index date. Calculations were performed in days, with 366 days included in 2020. Person-time at risk was evaluated in terms of person-weeks of follow-up and incident events of CVD and DM were identified in each four-week period. Data were then aggregated for the periods before the index date (‘Pre-Index’) and for four weeks after the index date (‘acute Covid-19’), 5 to 12 weeks after the index date (‘Post-acute Covid-19’) and from 13 to 52 weeks after the index date (‘Long Covid-19’). Incidence rates, with exact Poisson confidence intervals, were estimated for Covid-19 patients and controls. Poisson regression models were fitted, using the method of generalised estimating equations to allow for clustering on matched set, employing an exchangeable correlation structure. The effect of group (Covid-19 case or control), time (pre-index, 4 weeks from index, 5 to 12 weeks and 13 to 52 weeks) and the group-time interaction were estimated. The group-time interaction represented the net additional effect of acute-Covid, post-acute Covid or long Covid in Covid-19 cases, net of the baseline values for cases and the comparison with control participants. Analyses were adjusted for age, age-squared, gender, ethnicity, body mass index, systolic blood pressure, Charlson score, index month and index month squared. We conducted a sensitivity analysis including only cases with a positive polymerase chain reaction (PCR) test for SARS-CoV-19 infection. A logistic regression model was fitted to evaluate variables associated with PCR confirmation. All analyses were implemented in the R program, version 3.6.1.

## RESULTS

There were 516,985 Covid-19 cases, which were compared to a 516,985 controls matched on index date, practice, gender and age. After excluding matched sets that included either cases or controls with prevalent CVD or DM, there were 372,816 Covid-19 cases and 372,816 matched controls for further analysis. (Table 1) Covid-19 patients included 301,481 (80.9%) with clinical or laboratory confirmation of diagnosis on the index date, and 243,716 (65.4%) with a positive PCR test for SARS-CoV-2 on the index date. (Supplementary Table 1) Covid-19 cases and controls were similar with respect to matching variables of age and gender, there was a slight female predominance with median age of 33 years. Covid-19 cases were more likely to have defined values recorded for covariates. Covid-19 cases included slightly more of ‘Asian’ ethnicity, fewer current smokers, more who were overweight and obese, and more with comorbidity than controls, while blood pressure distributions appeared to be similar.

**Table 1:**
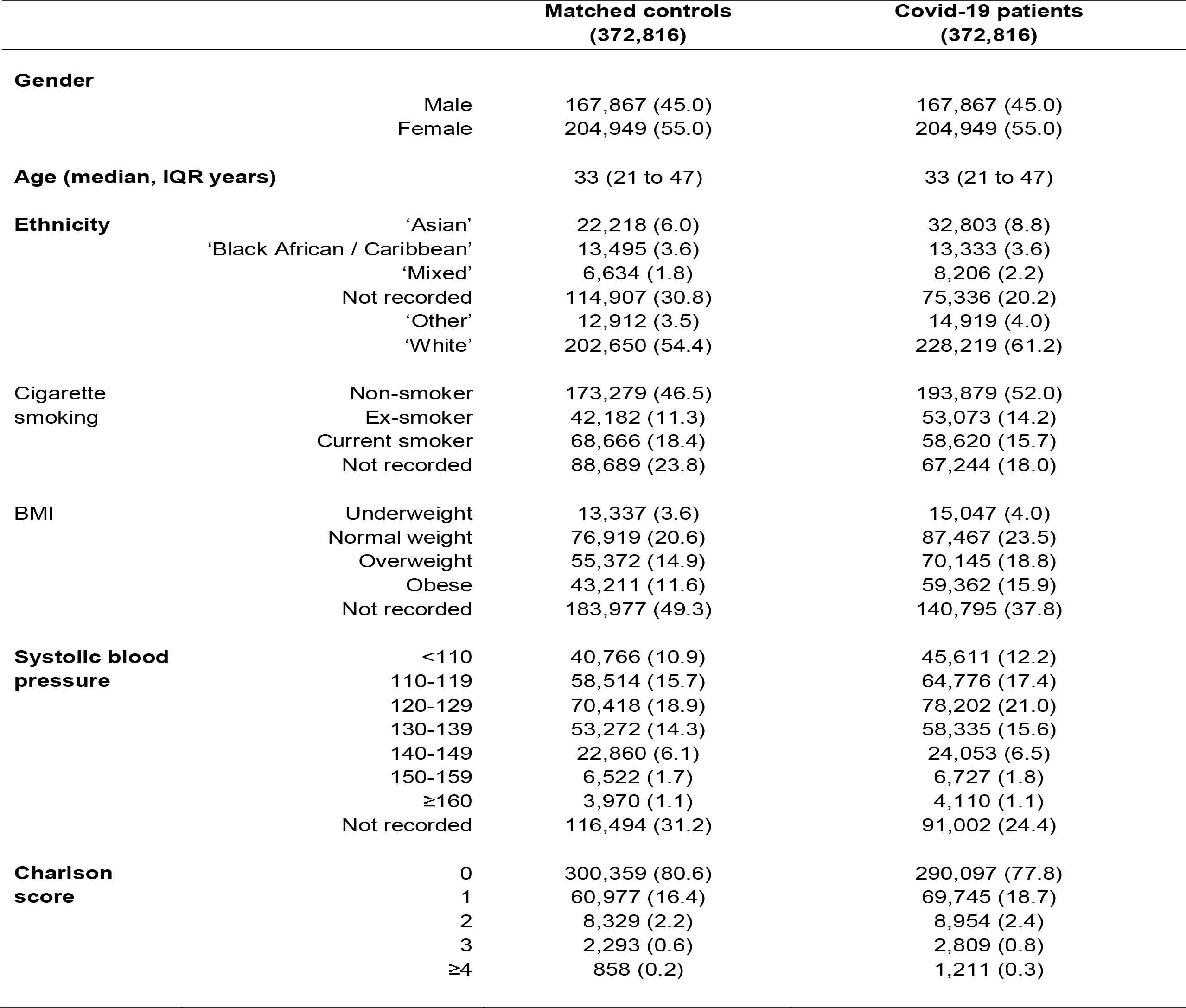
Characteristics of Covid-19 and matched control participants. Figures are frequencies (percent of column total).

Figure 1 shows the incidence of DM and CVD from 52 weeks before the index date to 52 weeks after for Covid-19 cases (red) and controls (blue). In the period before the index date, the incidence of diabetes was slightly higher in patients who went on to be diagnosed with Covid-19 than control participants; a similar difference was also observed for the incidence of cardiovascular diseases. In the first four weeks after Covid-19 diagnosis, there was a sharp increase in the incidence of diabetes and an even greater increase in cardiovascular diseases. The increase in CVD diagnoses was apparent in the last estimate for the pre-index period, which might imply that Covid-19 associated CVD might be recorded before Covid-19 diagnoses. In the remainder of the first year following Covid-19 diagnosis, the incidence of diabetes mellitus remained elevated for Covid-19 cases while CVD incidence declined to pre-morbid values.

**Figure 1:**
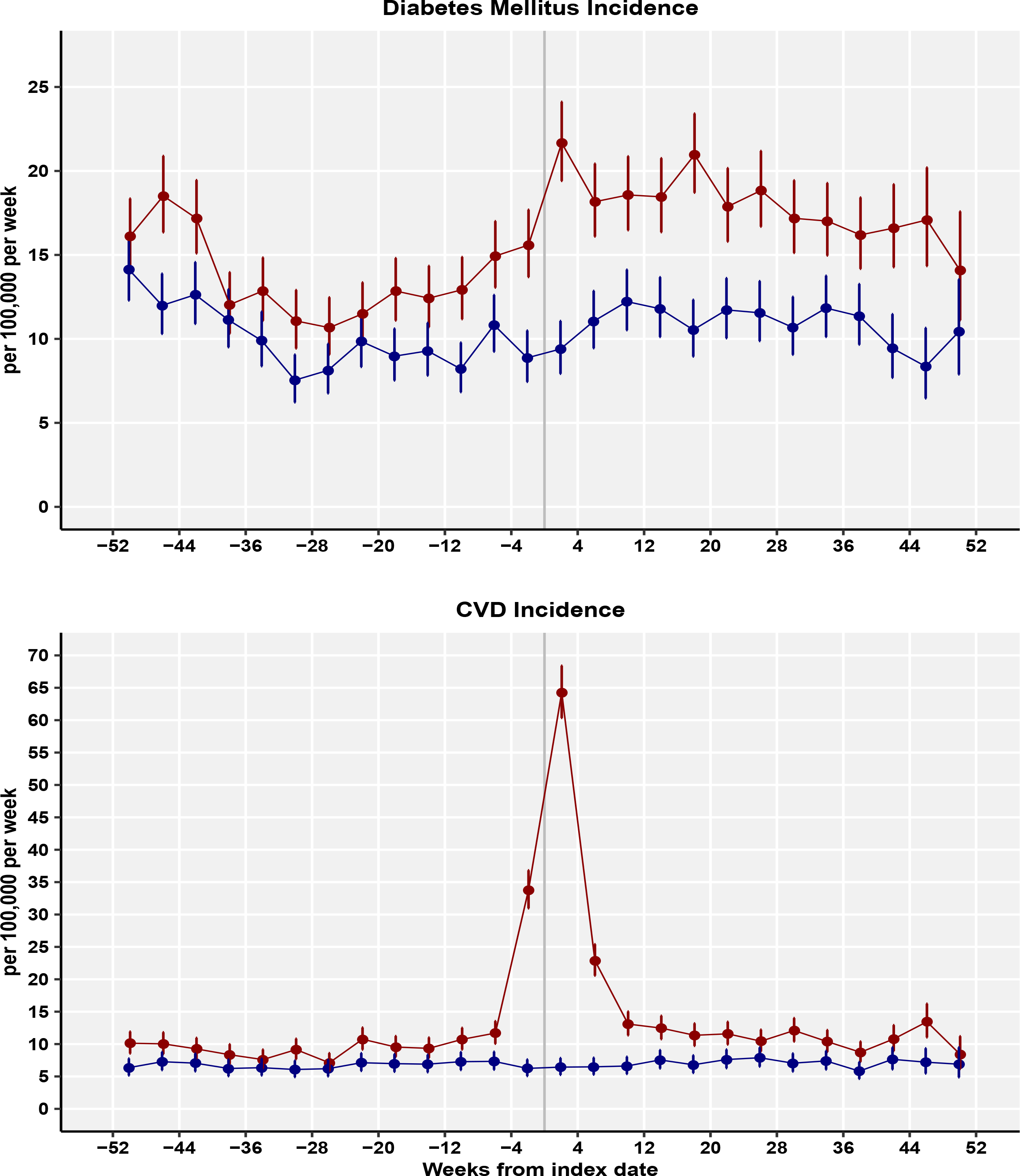
Incidence rates for diabetes mellitus and cardiovascular diseases (per 100,000 patient weeks) for Covid-19 patients and controls for 4-week periods.

Table 2 shows the incidence of DM and CVD aggregated into four periods: before the index date, within four weeks of the index date, from five to 12 weeks after the index date, and from 13 to 52 weeks after the index date. In the period before the index date, the incidence of diabetes was 13.73 (95% confidence interval 13.21 to 14.26) per 100,000 patient weeks in patients who were later diagnosed with Covid-19 but 10.10 (9.66 to 10.56) in controls who did not develop Covid-19; the incidence of CVD was 11.44 (10.96 to 11.93) per 100,000 patient weeks for cases and 6.72 (6.36 to 7.10) for controls respectively. During the first four weeks from Covid-19 diagnosis, the incidence of DM increased to 21.67 (19.40 to 24.12) and the incidence of CVD increased to 64.28 (60.33 to 68.42) per 100,000 patient weeks. In the post-acute and long Covid-19 periods, the incidence of CVD declined to 18.01 (16.53 to 19.58) and 11.04 (10.47 to 11.64) per 100,000 patient weeks respectively. Thus, in the long-Covid period overall, the incidence of CVD declined to pre-morbid levels. During the same periods, the incidence of DM remained elevated at 18.37 (16.88 to 19.96) per 100,000 in the post-acute period and 17.73 (17.00 to 18.48) per 100,000 in the long-Covid period.

**Table 2:** Mortality, CVD and DM events and rates by period for Covid-19 cases and controls. Figures are frequencies except where indicated.

| Phase | Case | Patient weeks | Deaths | Mortality rate per 100,000 patient weeks (95% CI) |  |  | CVD Events | CVD incidence per 100,000 patient weeks (95% CI) |  |  | Diabetes diagnoses | DM incidence per 100,000 patient weeks (95% CI) |  |  |
| --- | --- | --- | --- | --- | --- | --- | --- | --- | --- | --- | --- | --- | --- | --- |
| **Before index date** | Covid-19 | 19,028,859 | - | - | - | - | 2,177 | 11.44 | 10.96 | 11.93 | 2,612 | 13.73 | 13.21 | 14.26 |
|  | Controls | 19,499,049 | - | - | - | - | 1,311 | 6.72 | 6.36 | 7.10 | 1,970 | 10.10 | 9.66 | 10.56 |
| **Acute: up to 4 weeks from index** | Covid-19 | 1,537,024 | 1,260 | 81.98 | 77.51 | 86.63 | 988 | 64.28 | 60.33 | 68.42 | 333 | 21.67 | 19.40 | 24.12 |
|  | Controls | 1,521,612 | 55 | 3.61 | 2.72 | 4.70 | 98 | 6.44 | 5.23 | 7.85 | 143 | 9.40 | 7.92 | 11.07 |
| **Post-acute: 5 to 12 weeks** | Covid-19 | 3,037,595 | 442 | 14.55 | 13.23 | 15.97 | 547 | 18.01 | 16.53 | 19.58 | 558 | 18.37 | 16.88 | 19.96 |
|  | Controls | 3,008,270 | 104 | 3.46 | 2.82 | 4.19 | 197 | 6.55 | 5.67 | 7.53 | 350 | 11.63 | 10.45 | 12.92 |
| **Long: 13 to 52 weeks** | Covid-19 | 12,679,518 | 800 | 6.31 | 5.88 | 6.76 | 1,400 | 11.04 | 10.47 | 11.64 | 2,248 | 17.73 | 17.00 | 18.48 |
|  | Controls | 12,475,038 | 452 | 3.62 | 3.30 | 3.97 | 898 | 7.20 | 6.74 | 7.69 | 1,368 | 10.97 | 10.39 | 11.56 |

Figure 2 shows the incidence of categories of cardiovascular disease from 52 weeks before the Covid-19 index date to 52 weeks after. Pulmonary embolism showed the greatest increase during the acute Covid-19 period, with 29.93 cases per 100,000 patient weeks. The value was approximately three times higher than for atrial arrhythmias (9.95 per 100,000 per week) or venous thrombosis (8.65 per 100,000 per week); five times higher than for myocardial infarction and ischaemic heart disease (5.66 per 100,000 per week) or stroke (5.33 per 100,000 per week); and nearly than ten-times higher than the increase in heart failure (2.93 per 100,000 per week) or cardiomyopathy and myocarditis (1.56 per 100,000). While the relative effects differed for categories of CVD, the time course of changes in incidence was similar for each category of cardiovascular disease.

**Figure 2:**
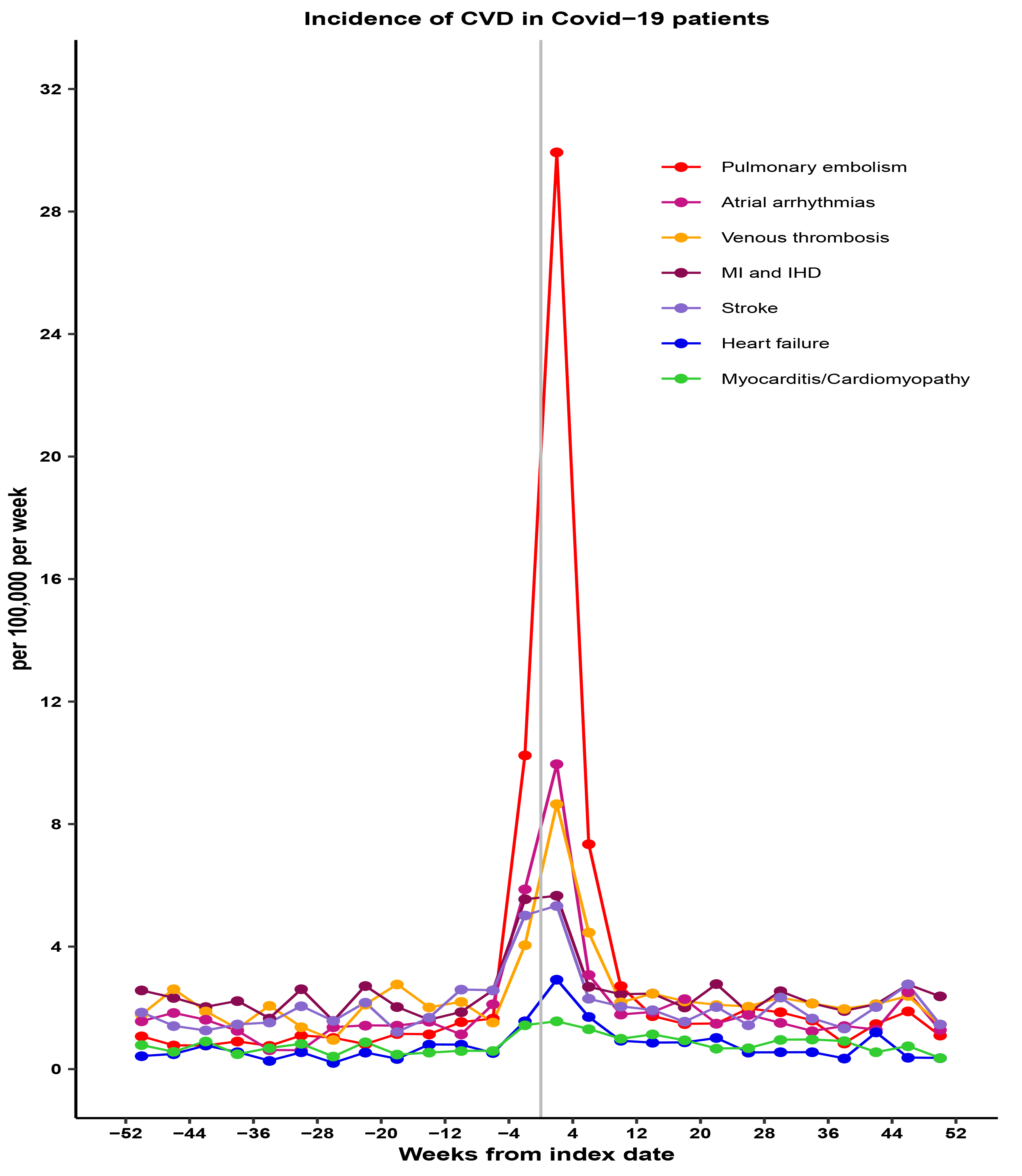
Incidence rates for categories of cardiovascular disease (per 100,000 patient weeks) for Covid-19 patients by 4-week periods.

Table 3 presents estimated incidence rate ratios adjusted for age, gender, ethnicity, smoking, body mass index, systolic blood pressure and Charlson comorbidity score. After allowing for underlying differences between cases and controls, and between the baseline pre-index period and follow-up, the net effect of acute Covid-19 on CVD incidence was estimated to be an adjusted rate ratio of 6.02 (95% confidence interval 4.84 to 7.47), while for DM incidence the adjusted rate ratio was 1.71 (1.40 to 2.10). In the post-acute Covid-19 period, the adjusted incidence rate ratio for CVD declined to 1.68 (1.41 to 2.01), while for DM in was 1.17 (1.01 to 1.36). In the long Covid-19 period, the adjusted incidence rate ratio for CVD was 0.95 (0.85 to 1.06) while for DM this remained elevated at 1.20 (1.09 to 1.31). In control participants, the incidence of CVD showed no overall change following the index date but there was evidence that the incidence of DM in controls was slightly higher for periods more than 4 weeks after the index date when compared with the pre-index period (Supplementary Table 2).

**Table 3:** Results of difference-in-difference analysis showing net effect of Covid-19 over baseline rate and comparison with controls. CI, confidence interval; RR, adjusted incidence rate ratio. (Estimates were adjusted for age, ethnicity, smoking, BMI category, SBP category, Charlson score, index month and matched set).

|  | Acute Covid-19 |  | Post-acute Covid-19 |  | Long Covid-19 |  |
| --- | --- | --- | --- | --- | --- | --- |
|  | RR (95% CI) | P value | RR (95% CI) | P value | RR (95% CI) | P value |
| **Diabetes mellitus** | 1.71 (1.40 to 2.10) | <0.001 | 1.17 (1.01 to 1.36) | 0.035 | 1.20 (1.09 to 1.31) | <0.001 |
| **All CVD outcomes** | 6.02 (4.84 to 7.47) | <0.001 | 1.68 (1.41 to 2.01) | <0.001 | 0.95 (0.85 to 1.06) | 0.377 |
| Pulmonary embolism | 14.54 (7.72 to 27.37) | <0.001 | 2.44 (1.49 to 4.02) | <0.001 | 0.70 (0.50 to 0.97) | 0.031 |
| Atrial arrhythmias | 6.58 (3.78 to 11.44) | <0.001 | 1.89 (1.17 to 3.06) | 0.010 | 0.88 (0.67 to 1.15) | 0.349 |
| Venous thrombosis | 5.44 (3.22 to 9.17) | <0.001 | 1.61 (1.09 to 2.37) | 0.017 | 1.07 (0.83 to 1.37) | 0.604 |
| Myocardial infarction and IHD | 3.53 (2.04 to 6.10) | <0.001 | 1.25 (0.83 to 1.90) | 0.288 | 1.10 (0.86 to 1.41) | 0.453 |
| Heart failure | 3.52 (1.51 to 8.20) | 0.004 | 1.75 (0.87 to 3.51) | 0.117 | 1.38 (0.84 to 2.26) | 0.205 |
| Stroke | 2.16 (1.35 to 3.44) | 0.001 | 0.99 (0.65 to 1.49) | 0.953 | 0.86 (0.67 to 1.11) | 0.236 |
| Cardiomyopathy and myocarditis | 1.91 (0.79 to 4.60) | 0.150 | 3.26 (1.30 to 8.15) | 0.012 | 0.97 (0.63 to 1.50) | 0.889 |

When categories of CVD were analysed separately (Table 3), pulmonary embolism (RR 14.5, 7.72 to 27.4), atrial arrythmias (6.58, 3.78 to 11.4) and venous thromboses (5.44, 3.22 to 9.17) showed the greatest relative increases in acute Covid-19 but myocardial infarction (3.53, 2.04 to 6.10), heart failure (3.52, 1.51 to 8.20) and stroke (2.16, 1.35 to 3.44) were also increased. These conditions declined substantially in the post-acute Covid-19 period. There was some evidence that cardiomyopathy and myocarditis might be more frequent in the post-acute period (3.26, 1.30 to 8.15) than in the acute illness. There was no evidence that any of these categories of CVD showed a net increase during the long-Covid-19 period.

The median age at diagnosis of DM was 45 (interquartile range 33 to 55) years for Covid-19 cases and 44 (31 to 55) years for controls; for CVD, the median age at diagnosis was 57 (47 to 67) for Covid-19 cases and 57 (46 to 68) for controls. (Supplementary Table 3) The distribution of gender, ethnicity, smoking and obesity were generally consistent across stages of Covid-19 infection. Patients diagnosed with diabetes in the context of acute Covid-19 were more likely to be prescribed insulin with 91 days of diagnosis (45/333, 13.5%, compared with 10/143, 7.0%, for controls) but there was no clear evidence of an increase in Type 1 DM at any stage of the illness. (Supplementary Table 3) There were 5,671 (1.5%) control patients and 14,783 (4.0%) Covid-19 patients who received one or more primary care prescriptions for oral, enteral, intramuscular or intravenous glucocorticoids from four weeks before the index date to the end of follow-up. There were 86 new diagnoses of diabetes in glucocorticoid-treated controls, and 280 in glucocorticoid-treated cases during the same period.

### Sensitivity analysis

There were 243,716 Covid-19 cases with a positive PCR test recorded on the index date. Compared with the entire sample, patients with PCR test confirmation of Covid-19 tended to be younger, were more often white, male, non-smokers, with normal body mass index. The odds of PCR confirmation decreased by 2.3% per year increase in age. Results of the sensitivity analysis are shown in Supplementary Table 4. Although rates of CVD and DM were lower in the sample with PCR confirmed Covid-19 infection, consistent with their generally lower cardiometabolic risk profile, differences between Covid-19 cases and controls, and changes over time following Covid-19 infection, were consistent with those observed in the whole sample.

## DISCUSSION

### Main findings

Drawing on longitudinal electronic health records from primary care, this study compared Covid-19 patients with matched population controls from one year before a Covid-19 diagnosis to one year after. The study showed that patients recorded with Covid-19 in primary care are at slightly greater risk of CVD and DM even before the Covid-19 diagnosis. These elevated risks were associated with higher body mass index, greater comorbidity and a higher proportion of ‘Asian’ ethnicity. After allowing for baseline differences between Covid-19 patients and controls, new DM diagnoses increased 1.7-fold during acute Covid-19 and remained elevated during both post-acute and long Covid-19. There was evidence of increased glucocorticoid exposure in Covid-19 patients, but this might be associated with only a very small proportion of new DM events. New CVD events were increased more than six-fold during acute Covid-19 and more than 50% in post-acute Covid-19. This increase was largely driven by pulmonary embolism diagnoses, but atrial arrythmias, venous thrombosis, myocardial infarction, stroke and heart failure also showed more modest increases. Cardiomyopathy and myocarditis only showed evidence of increase in the post-Covid-19 period, which might perhaps be explained by the lower specificity of associated symptoms and delayed recognition. CVD events returned to baseline levels during the long Covid-19 period. Control patients showed no change in CVD events but there was a slight increase in DM events following the index date.

### Comparison with other studies

The long-term outcomes of Covid-19 infection represent an emerging area of research and public concern, but definitions are not universally agreed. This study drew on clinical recommendations that distinguish ‘acute’ (four weeks), ‘post-acute’ (three months) and ‘long’ (one year) periods after Covid-19 infection.^17^ ‘Long Covid’ is often self-identified by patients with the concept focusing on symptom burden and impacts on health-related quality of life.^17,22^ These subjective outcomes of Covid-19 infection may be heterogenous in nature and their occurrence and persistence may depend on patient characteristics including age, gender and comorbidity, as well as the severity of the initial Covid-19 illness and intensity and duration of therapeutic support in the acute illness.^22,23^ It is well established that pre-existing hypertension and ischaemic heart disease are associated with more severe illness and greater mortality in acute Covid-19 infection.^24^ Hospitalised patients may experience a range of cardiovascular complications including arrhythmias, heart failure and thrombotic complications.^24^ However, there are few studies with long-term follow-up of patients without pre-existing cardiovascular disease. In a pre-print, Knight et al.^25^ reported on cardiovascular outcomes of a large population in England. Their analyses suggested that CVD outcomes might remain increased for up to 49 weeks following Covid-19 infection.^25^ However, the analyses did not include a baseline period before the Covid-19 infection that might have provided more precise evaluation of baseline differences in risk between Covid-19 cases and controls.

Pre-existing diabetes mellitus is also associated with more severe illness from Covid-19^26^ but some studies suggest that Covid-19 may be associated with new onset diabetes. A systematic review of eight reports of patients hospitalised in the early stages of the Covid-19 pandemic found that 14.4% of patients developed new onset diabetes following the illness.^27^ A possible effect of SARS-CoV-2 infection on pancreatic function is suggested by the finding that the virus infects pancreatic beta cells,^28^ reduces insulin production and promotes beta-cell apoptosis.^29^ However, Covid-19 may also lead to reduced physical activity and deconditioning^30^ leading to greater insulin resistance. Contacts with medical care may also lead to increased opportunities to detect previously undiagnosed diabetes. Previous studies have often reported hospital-based cohorts, with smaller samples or shorter durations of follow-up. This large population-based study shows that patients diagnosed with Covid-19 have a slightly higher baseline risk of diabetes mellitus. There is evidence that new-onset diabetes is increased during the acute Covid-19 illness and there is evidence that a net increase in diabetes persists for at least 12 months following the Covid-19 illness. Analysis of electronic records does not permit unequivocal distinction of type I and type II diabetes phenotypes. There was evidence that patients diagnosed during the acute illness might be more likely to be prescribed insulin but there was no evidence for increased type I diabetes overall, nor was there evidence that glucocorticoid exposure might account for a high proportion of cases.

### Strengths and limitations

This study drew on a large, longitudinal population-based data resource that enabled us to conduct a matched analysis of mortality and new CVD and DM diagnoses for up to one year following Covid-19.[28] The study drew on clinical records with several limitations. We included patients with both confirmed and suspected Covid-19, but a sensitivity analysis found that restricting the analysis to PCR confirmed infections would not alter conclusions. However, PCR testing was associated with patient characteristics and reliance on PCR confirmation for participant selection might lead to bias. The database enabled adjustment for a range of important covariates, but these were not always completely recorded. This may reflect that this was a relatively young to middle-aged cohort lacking in long-term conditions that would warrant regular consultations and monitoring. In health records, values are commonly missing ‘not at random’ making the application of imputation methods more difficult. We did not include a measure of deprivation, which is associated both with diabetes and cardiovascular disease, but cases and controls were matched for family practice, providing partial control for area-based measures including deprivation. Analyses did not include measures of the severity of illness in Covid-19. However, the concept of ‘severity’ might be difficult to operationalise in a study of Covid-19 complications because a greater number of complications might be indicative of more severe illness. We did not have access to data for secondary care prescribing, consequently exposure to glucocorticoids might be under-estimated.^18^ The observational nature of the study limits causal inferences concerning whether the increased risk of new CVD and DM diagnoses may result from Covid-19, or whether undiagnosed CVD and DM were more prevalent among Covid-19 cases, or whether Covid-19 aggravated or altered the natural history of pre-existing disease.

### Conclusion

This study provides evidence that DM incidence remains elevated for up to one year following Covid-19 infection. Future studies should aim to characterise the phenotypic characteristics and underlying mechanisms of Covid-19 associated diabetes mellitus. Advice to patients recovering from Covid-19 should include measures to reduce diabetes risk, including diet, weight management and physical activity levels. CVD is increased early after Covid-19 infection, mainly from pulmonary embolism, atrial arrhythmias and venous thromboses, and the risk is increased for up to three months. Future research should determine which patients are at greatest risk, and whether these cardiovascular complications may be prevented through prophylactic measures.

## Data Availability

All data produced in the present study are available upon reasonable request to the authors. The study is based on data from the Clinical Practice Research Datalink (CPRD) obtained under license from the UK Medicines and Healthcare Products Regulatory Agency (MHRA); however, the interpretation and conclusions contained in this report are those of the authors alone. Requests for access to data from the study should be addressed to the corresponding author at. All proposals requesting data access will require approval from CPRD before data release.

## Ethical approval

The protocol was given scientific and ethical approval by the CPRD Independent Scientific Advisory Committee (ISAC protocol 20_00265R). CPRD holds over-arching research ethics committee approval for the conduct of research using fully anonymised electronic health records data from the CPRD databases.

## Study protocol

The study protocol has been published in summary form by the CPRD/MHRA and can be accessed <u>here</u>.

## Contributors

ERP conceived the idea for study. ERP and MG analysed the data with advice from AD, XS, PC and AMS. ERP and MG drafted the paper. All authors reviewed the analysis, contributed to drafts of the paper and approved the final draft. MG is guarantor.

## Data sharing

The study is based on data from the Clinical Practice Research Datalink (CPRD) obtained under license from the UK Medicines and Healthcare Products Regulatory Agency (MHRA); however, the interpretation and conclusions contained in this report are those of the authors alone. Requests for access to data from the study should be addressed to the corresponding author at. All proposals requesting data access will require approval from CPRD before data release.

