## Supplementary Tables for "Differential impact of Covid-19 on incidence of diabetes mellitus and cardiovascular diseases in acute, post-acute and long Covid-19: population-based cohort study in the United Kingdom"

**Supplementary Table 1: Medical codes associated with Covid-19 index date in 385,349 Covid-19 cases**.

| **Medical Term** | **Freq.** |
| --- | --- |
| SARS-CoV-2 (severe acute respiratory syndrome coronavirus 2) RNA detection result positive | 242,289 |
| SARS-CoV-2 (severe acute respiratory syndrome coronavirus 2) detection result positive | 16,375 |
| COVID-19 confirmed by laboratory test | 16,208 |
| COVID-19 | 6,755 |
| SARS-CoV-2 (severe acute respiratory syndrome coronavirus 2) IgG detection result positive | 4,968 |
| SARS-CoV-2 (severe acute respiratory syndrome coronavirus 2) antigen detection result positive | 2,586 |
| COVID-19 | 2,363 |
| Coronavirus infection | 1,938 |
| Disease caused by 2019-nCoV | 1,654 |
| Disease caused by 2019-nCoV (novel coronavirus) | 1,158 |
| COVID-19 severity score | 866 |
| Detection of 2019-nCoV (novel coronavirus) using polymerase chain reaction technique | 852 |
| Detection of SARS-CoV-2 using polymerase chain reaction | 575 |
| Confirmed 2019-nCoV (novel coronavirus) infection | 554 |
| COVID-19 confirmed using clinical diagnostic criteria | 524 |
| SARS-CoV-2 (severe acute respiratory syndrome coronavirus 2) detected | 359 |
| Assessment using COVID-19 severity scale | 347 |
| SARS-CoV-2 (severe acute respiratory syndrome coronavirus 2) antibody detection result positive | 259 |
| SARS-CoV-2 (severe acute respiratory syndrome coronavirus 2) IgA detection result positive | 257 |
| Pneumonia caused by SARS-CoV-2 (severe acute respiratory syndrome coronavirus 2) | 162 |
| COVID-19 severity scale | 104 |
| SARS-CoV-2 (severe acute respiratory syndrome coronavirus 2) IgM detection result positive | 80 |
| COVID-19 confirmed clinically | 52 |
| Possible COVID-19 | 47 |
| Pneumonia caused by SARS-CoV-2 (severe acute respiratory syndrome coronavirus 2) | 35 |
| Pneumonia caused by 2019-nCoV (novel coronavirus) | 18 |
| Pneumonia caused by 2019-nCoV (novel coronavirus) | 17 |
| COVID-19 pneumonia | 16 |
| Probable COVID-19 confirmed using clinical diagnostic criteria | 12 |
| Other codes (15 codes) | 51 |
| **‘Suspected’ Covid-19** |  |
| Suspected COVID-19 | 29,558 |
| Telephone consultation for suspected 2019-nCoV (novel coronavirus) | 15,707 |
| Suspected disease caused by 2019-nCoV (novel coronavirus) | 13,231 |
| Suspected coronavirus infection | 9,139 |
| Suspected 2019-nCoV (novel coronavirus) infection | 3,076 |
| Telephone consultation for suspected SARS-CoV-2 | 520 |
| Suspected disease caused by Wuhan 2019-nCoV (novel coronavirus) | 95 |
| Other codes (3 codes) | 9 |

**Supplementary Table 2: Results of Poisson regression models showing main effects of group and time period and group by time interaction. CI, confidence interval; RR, adjusted incidence rate ratio. (Estimates were adjusted for age, ethnicity, smoking, BMI category, SBP category, Charlson score, index month and matched set).**

|  | **Incidence of CVD** | | **Incidence of DM** |  |
| --- | --- | --- | --- | --- |
|  | **RR (95% CI)** | **P value** | **RR (95% CI)** | **P value** |
| **Net effect of Covid-19^a,b^** |  |  |  |  |
| Acute Covid-19 (up to 4 weeks from index) | 6.02 (4.84 to 7.47) | <0.001 | 1.71 (1.40 to 2.10) | <0.001 |
| Post-acute Covid-19 (5-12 weeks from index) | 1.68 (1.41 to 2.01) | <0.001 | 1.17 (1.01 to 1.36) | 0.035 |
| Long Covid-19 (13-52 weeks from index) | 0.95 (0.85 to 1.06) | 0.377 | 1.20 (1.09 to 1.31) | <0.001 |
| **Controls^c^** |  |  |  |  |
| Before index date | Ref. |  | Ref. |  |
| 4 weeks from index date | 0.97 (0.79 to 1.19) | 0.776 | 0.94 (0.79 to 1.11) | 0.466 |
| 5-12 weeks from index date | 0.99 (0.85 to 1.15) | 0.868 | 1.16 (1.04 to 1.31) | 0.010 |
| 13-52 weeks from index date | 1.05 (0.96 to 1.15) | 0.269 | 1.09 (1.02 to 1.17) | 0.016 |
| **Overall difference between Covid-19 patients and controls^d^** | |  |  |  |
| Control | Ref. |  | Ref. |  |
| Covid-19 patients | 1.58 (1.47 to 1.69) | <0.001 | 1.14 (1.08 to 1.21) | <0.001 |

^a^group by time interaction; ^b^ additional effect, net of rate in pre-index period for cases and rate in controls in same period; ^c^ ‘time’ effect; ^d^ ‘group’ effect;

**Supplementary Table 3: Characteristics of case and control patients diagnosed with DM or CVD during follow-up.**

|  |  | **Pre-Index** | **Acute** | **Post-Acute** | **Long** |
| --- | --- | --- | --- | --- | --- |
| **Incident diabetes mellitus patients (9,582)** | | |  |  |  |
| Number | Covid-19 | 2,612 | 333 | 558 | 2,248 |
|  | Controls | 1,970 | 143 | 350 | 1,368 |
| Age (mean, years) | Covid-19 | 44 (33 to 55) | 48 (37 to 57) | 45 (34 to 55) | 44 (33 to 55) |
|  | Controls | 44 (31 to 55) | 40 (30 to 55) | 43 (31 to 54) | 44 (31 to 54) |
| Age≤35 years | Covid-19 | 778 (29.8) | 71 (21.3) | 168 (30.1) | 702 (31.2) |
|  | Controls | 660 (33.5) | 55 (38.5) | 123 (35.1) | 472 (34.5) |
| Insulin within 91 days of diagnosis | Covid-19 | 110 (4.2) | 45 (13.5) | 42 (7.5) | 79 (3.5) |
|  | Controls | 83 (4.2) | 10 (7.0) | 19 (5.4) | 71 (5.2) |
| ‘Type 1 DM’ | Covid-19 | 55 (2.1) | 11 (3.3) | 21 (3.8) | 44 (2.0) |
|  | Controls | 53 (2.7) | 8 (5.6) | 14 (4.0) | 43 (3.1) |
| Male | Covid-19 | 1,046 (40.0) | 179 (53.8) | 243 (43.5) | 809 (36.0) |
|  | Controls | 657 (33.4) | 46 (32.2) | 118 (33.7) | 375 (34.7) |
| Current smoker | Covid-19 | 500 (19.1) | 54 (16.2) | 103 (18.5) | 430 (19.1) |
|  | Controls | 446 (22.6) | 36 (25.1) | 86 (24.5) | 315 (23.0) |
| Obese | Covid-19 | 1,165 (44.6) | 142 (42.6) | 210 (37.6) | 934 (41.5) |
|  | Controls | 810 (41.1) | 56 (39.2) | 121 (34.6) | 498 (36.4) |
| ‘Asian’ ethnicity | Covid-19 | 413 (15.8) | 60 (18.0) | 83 (14.9) | 331 (14.7) |
|  | Controls | 202 (10.3) | 19 (13.3) | 33 (9.4) | 161 (11.8) |
| **Incident CVD patients (7,616)** | |  |  |  |  |
| Number | Covid-19 | 2,177 | 988 | 547 | 1,400 |
|  | Controls | 1,311 | 98 | 197 | 898 |
| Age (median, IQR years) | Covid-19 | 57 (48 to 69) | 58 (48 to 68) | 57 (47 to 66) | 55 (44 to 64) |
|  | Controls | 57 (46 to 68) | 56 (46 to 68) | 56 (46 to 64) | 57 46 to 68) |
| Male | Covid-19 | 1,094 (50.3) | 583 (59.0) | 547 (55.4) | 655 (46.8) |
|  | Controls | 637 (48.6) | 54 (55.1) | 197 (47.2) | 448 (49.9) |
| Current smoker | Covid-19 | 438 (20.1) | 172 (17.4) | 116 (21.2) | 299 (21.4) |
|  | Controls | 297 (22.7) | 29 (29.6) | 54 (27.4) | 225 (25.1) |
| Obese | Covid-19 | 682 (31.3) | 342 (34.6) | 178 (32.5) | 419 (29.9) |
|  | Controls | 367 (28.0) | 28 (28.6) | 46 (23.4) | 220 (24.5) |
| ‘Asian’ ethnicity | Covid-19 | 101 (4.6) | 55 (5.6) | 36 (6.6) | 72 (5.1) |
|  | Controls | 47 (3.6) | 3 (3.1) | 4 (2.0) | 34 (3.8) |

**Supplementary Table 4: Results of a sensitivity analysis on 243,716 Covid-19 cases confirmed by polymerase chain reaction (PCR) test and matched controls. Figures are frequencies except where indicated.**

| **Phase** | **Case** | **Patient weeks** | **Deaths** | **Mortality rate per 100,000 patient weeks (95% CI)** | | | **CVD Events** | **CVD incidence per 100,000 patient weeks (95% CI)** | | | **Diabetes diagnoses** | **DM incidence per 100,000 patient weeks (95% CI)** | | |
| --- | --- | --- | --- | --- | --- | --- | --- | --- | --- | --- | --- | --- | --- | --- |
|  |  |  | **Events** | **Rate** | **Lower** | **Upper** | **Events** | **Rate** | **Lower** | **Upper** | **Events** | **Rate** | **Lower** | **Upper** |
| **Before index date** | Covid-19 | 12,424,861 | - | - | - | 0.03 | 859 | 6.91 | 6.46 | 7.39 | 1376 | 11.07 | 10.50 | 11.68 |
|  | Controls | 12,723,759 | - | - | - | 0.03 | 739 | 5.81 | 5.40 | 6.24 | 1135 | 8.92 | 8.41 | 9.45 |
| **Acute: up to 4 weeks from index** | Covid-19 | 1,006,508 | 325 | 32.29 | 28.87 | 36.00 | 301 | 29.91 | 26.62 | 33.48 | 150 | 14.90 | 12.61 | 17.49 |
|  | Controls | 9,944,87.7 | 23 | 2.31 | 1.47 | 3.47 | 62 | 6.23 | 4.78 | 7.99 | 89 | 8.95 | 7.19 | 11.01 |
| **Post-acute: 5 to 12 weeks** | Covid-19 | 1,993,383 | 107 | 5.37 | 4.40 | 6.49 | 216 | 10.84 | 9.44 | 12.38 | 328 | 16.45 | 14.72 | 18.34 |
|  | Controls | 1,965,881 | 49 | 2.49 | 1.84 | 3.30 | 111 | 5.65 | 4.64 | 6.80 | 234 | 11.90 | 10.43 | 13.53 |
| **Long: 13 to 52 weeks** | Covid-19 | 8,058,223 | 204 | 2.53 | 2.20 | 2.90 | 595 | 7.38 | 6.80 | 8.00 | 1233 | 15.30 | 14.46 | 16.18 |
|  | Controls | 7,871,591 | 226 | 2.87 | 2.51 | 3.27 | 510 | 6.48 | 5.93 | 7.07 | 862 | 10.95 | 10.23 | 11.71 |
